# COVID-19 INDUCES SENESCENCE AND EXHAUSTION OF T CELLS IN PATIENTS WITH MILD/MODERATE AND SEVERE DISEASE DURING A SEVEN-DAY INTERVAL

**DOI:** 10.1101/2023.01.16.23284612

**Authors:** Rodrigo Balsinha Pedroso, Lucas Haniel Araújo Ventura, Lícia Torres, Giovanna Caliman Camatta, Felipe Caixeta, Leandro Souza Nascimento, Catarina Mota, Ana Catarina Mendes, Filipa Ribeiro, Henrique Cerqueira Guimarães, Rafael Calvão Barbuto, Gabriela Silveira-Nunes, Andrea Teixeira-Carvalho, Luis Graça, Ana Maria Caetano Faria

## Abstract

Risk factors for the development of severe COVID-19 include several comorbidities, but age was the most striking one since elderly people were disproportionately affected by SARS-Cov-2. Major drivers that can explain this markedly unfavourable response in the elderly are inflammaging and immunosenescence. Recent reports have shown that the relationship between immunosenescence and COVID-19 can be bidirectional, since hospitalized patients with severe COVID-19 have an accumulation of senescent T cells suggesting that immunosenescence can be also exacerbated by SARS-CoV-2 infection. Therefore, the present work was designed to examine the emergence of immunosenescence in a longitudinal study in two distinct cohorts of COVID-19 patients, and to determine whether the senescence alterations were restricted to severe cases of the disease. Our data, with patients from Portugal and Brazil, identified their distinctive inflammatory profile and provided evidence of increased frequencies of senescent and exhausted T cells within a seven-day period in patients with mild to severe COVID-19. These results support the view that SARS-CoV2 infection can accelerate immunosenescence in both CD4 and CD8 T cell compartments in a short period of time.

## Introduction

Since 2019 when it first started in China, the Corona Virus Disease-2019 (COVID-19) pandemic caused by the severe acute respiratory syndrome coronavirus 2 (SARS-Cov-2) has been responsible for the death of more than 6 million people worldwide (WHO, 2020a). Clinical manifestations of COVID-19 range from asymptomatic to severe pneumonia associated with extensive immune-inflammatory response, a clinical condition that can lead to multiorgan failure and prolonged complications (Osuchowski et al, 2021).

Risk factors for the development of severe COVID-19 include obesity, diabetes, high blood pressure, male sex, and age, with the latter being the most striking, since elderly people were disproportionately affected by SARS-Cov-2 (Li et al, 2020; Guo et al, 2020; Rahman & Sathi, 2020; Xu & Chen, 2020; Wu et al, 2020; Zhou et al, 2020). Among the reasons for this markedly unfavourable response in the elderly, immunosenescence and *inflammaging* are major drivers of this outcome (Mueller, McNamara, Sinclair, 2020; Camell et al, 2021). Immunosenescence is the aging of the immune system and is mainly characterized by a decrease in naïve T cell numbers together with an accumulation of CD4^+^ and CD8^+^ memory and terminal effector T cells, resulting in increased vulnerability to infections and impaired response to vaccination (Hassoueneh et al, 2016; Fulop et al, 2018; Aiello et al, 2019). Moreover, aging T lymphocytes like other senescent cells tend to develop a secretory-associated senescent phenotype (SASP), characterized by the production and secretion of IL-6, IL-1, IL-8, IL-18, and TNF along with other inflammatory mediators. Senescent cell accumulation as well as other aging-related disturbances (mitochondrial dysfunction, dysbiosis of microbiota, innate immune responses) lead to a chronic low grade inflammatory state known as *inflammaging* (Fulop et al, 2018; Batista et al, 2020; Heath and Grant, 2020). Continuous immune dysregulation induced by prolonged exposure to infectious and non-infectious antigens can also drive immune cell aging, since the accumulation of inflammatory mediators secreted in response to consecutive infections further contributes for the impairment of the adaptive immune response, boosting immunossenescence and *inflammaging* (Fulop et al, 2018; Goronzy & Weyand, 2019; Elyahu & Monsonego, 2021). Chronic infections, including those caused by the human cytomegalovirus (CMV), HIV, hepatitis B virus (HBV) and human papillomavirus (HPV), are sources of constant stimulation to the immune system, resulting in the accumulation of terminally differentiated senescent T cell populations and, consequently, immunesenescence (Solana et al, 2012; Fulop, Larbi, Pawelec, 2013). Similarly, although it is not a persistent infection, SARS-Cov-2 can also trigger the accumulation of exhausted and senescent T cell phenotypes due to its extensive activation and clonal expansion (Diao et al, 2020; Zheng et al, 2020b). Recent publications have shown that severe COVID-19 patients presented an increased frequency of terminally differentiated CD4 and CD8 T cells when compared to controls matched for sex and age (Zheng et al, 2020a; Arcanjo et al, 2021). However, there is no longitudinal study clearly demonstrating that SARS-CoV-2 infection leads to accelerated immunosenescence in distinct populations and in different forms of COVID-19.

In this study, we investigated the evolution of the inflammatory response and immunosenescence of COVID-19 patients from Belo Horizonte, Brazil, and Lisbon, Portugal. Blood samples were collected during ambulatory care or hospitalization, and one week later, allowing the evaluation of the impact of the disease over a period of seven days. Our results showed that SARS-CoV-2 infection induces T cell senescence in both populations in a short period of time, in both moderate and severe cases, with this effect being more prominent in the latter.

## Materials and Methods

### Ethics statement

The research protocol was approved by the National Research Ethics Commission (CONEP # 5.190.260) of Brazil and by the Lisbon Academic Medical Center Ethics Committee (ref. no. 306/20). The study was conducted in accordance with the Helsinki Declaration for research involving humans. All participants, including healthy controls, agreed to participate voluntarily in this study without financial support and signed an informed consent.

### Study design

This longitudinal study was performed at two healthcare centres: Centro Hospitalar Universitário Lisboa Norte (CHULN) in Lisbon, Portugal, and Hospital Risoleta Tolentino Neves in Belo Horizonte, Brazil. Non-vaccinated patients were recruited within the first three days of hospitalization or while in ambulatory care, after presenting a positive nasopharyngeal swab test for SARS-CoV-2 by real-time reverse transcriptase-polymerase chain reaction (RT-PCR) performed upon admission at the health care units. A total of 20 infected patients from Lisbon and 10 from Belo Horizonte agreed to participate in this study. For the longitudinal follow-up, blood samples were collected at the time of enrolment (T0) and seven days later (T7). Each city had a control group formed according to the following criteria: testing negative for COVID-19 in the RT-PCR test and not having presented any symptom the week before blood sample collection. Individuals with prior inflammatory diseases, such as cancer and renal failure were excluded. Data collection was carried out between August and October 2020 in Lisbon and June and August in Belo Horizonte.

Disease severity classification was based on the WHO criteria (WHO, 2020b). Mild cases were considered asymptomatic or those treated in an outpatient clinic. Moderate cases included hospitalized patients without oxygen support or using a low to moderate flow oxygen by mask or nasal prongs. Severe cases were characterized as the need for high flow oxygen, mechanical ventilation, vasopressors, dialysis, or extracorporeal membrane oxygenation (ECMO) (WHO, 2020b).

### Blood collection and isolation of PBMCs

Peripheral blood mononuclear cells (PBMCs) were obtained by blood collection from COVID-19 patients and healthy controls using heparinized vacuette tubes. The purification was attained by Ficoll gradient (Histopaque-1077; Sigma, cat #10771) centrifugation at room temperature, in a 1:2 ratio, for 40 minutes, at 600xg and without brake-induced end-centrifugation deceleration phase. After that, the PBMCs were collected and washed with Roswell Park Memorial Institute (RPMI) 1640 basal medium. Red blood cells were lysed using a lysis buffer. Cells were counted and stored in fetal bovine serum supplemented with 10% dimethyl sulfoxide at -80 °C until further use. Blood serum was also obtained by centrifuging whole blood collected in sera tubes for 10 minutes at 500xg.

### Immunophenotype by polychromatic flow cytometry

PBMCs (1×10^6^) were first stained with LIVE/DEAD fixable aqua dead cell stain (ThermoFisher Scientifc, cat #L34957) and antibodies fluorophore-conjugated human to surface markers. anti-CD8 (RPA-T8, cat# 562282), -CD4 (SK3, cat# 557852), -CD25 (M-A251, cat# 555432), -CD278 (ICOS; DX29, cat# 562833), -CD57 (NK-1, cat# 563896), -CD279 (PD-1; EH12.1, cat# 563245), -TIGIT (741182, cat# 747840), -CD28 (CD28.2, cat# 561368), -CD45RO (cat# 304234), -CD3 (cat# 317340), -CCR7 (cat# 353216) and -KLRG1 (cat# 138427).

After the surface staining, cells were fixed and permeabilized using the Foxp3/Transcription Factor Staining Buffer Set from eBioscience (cat #00-5523-00) and stained for -Foxp3 from ImmunoTools (3G3, cat# 21276106). Single colors for fluorescence compensation were prepared with antibody capture compensation beads (BD Biosciences). Cell samples were acquired in a BD LSRFortessa cell analyzer (BD Biosciences) coupled to computers with DIVA and FlowJo-10 *software* (Tree Star).

### Luminex-Multiplex measurement of serum cytokines, chemokines, and growth factors

A Bio-Rad Laboratories kit (Bio-Plex Pro Human Cytokine Standard) was used to analyse multiple serum mediators simultaneously with the Bio-Plex 200 system from Bio-Rad, following storage and processing protocols standardized by the Laboratory of Biomarkers (IRR-FIOCRUZ/MG). Samples were transported and stored at a temperature of -80ºC. Analyzes were performed using Bioplex™ xPONENT version 3.1 software (Bio-Rad) and included the following panel of analytes: IL-1β, IL-1ra, IL-2, IL-4, IL-5, IL-6, CXCL-8 (IL-8), IL-10, IL-12p70, IL-13, GM-CSF, IFN-γ, CCL-2 (MCP-1) and TNF-α. The panel of biomarkers included in the kit and their standard settings is presented in Supplementary Table 1.

### Statistical analysis

Statistical analyses were performed using GraphPad Prism 8.0 *software* (GraphPad Software, San Diego, CA, USA). All data acquired from Luminex-Multiplex were converted to logarithmic scale to normalize the data, and a lognormality test was performed to confirm data normality. Wilcoxon test was applied in non-parametric data to compare samples of the same patient at T0 and T7. Student’s t-test was used for analysing parametric data.

Radar plots highlight the contribution of different cytokines to immunological profile. This analysis allows to convert quantitative cytokine measurements into a categorical variable of low and high cytokine producers as previously proposed by Luiza-Silva et al. (2011). Briefly, this approach categorizes each subject as “Low” or “High” cytokine producer, taking the global median value as a specific cut-off edge for each cytokine. For the calculation of the global median the whole universe of data obtained for the groups was considered. Following data categorization, the frequency of “High Cytokine Producers” was calculated for each group. On radar charts, each axis represents the percentage (%) of volunteers categorized as “High Cytokine Producers” in each group. Then, connecting the values of each axis forms a central polygonal area that represents the immunological profile. Relevant productions were considered when the percentage of “High Cytokine Producers” of a given cytokine was greater than 50%.

The correlation matrix was built using the cor() function of the statistical analysis package stats v3.6.2 available in the R v4.0 environment. Spearman’s correlation was used considering the non-parametric nature of the data, considering only correlations with p<0.05.

Statistical analysis of data on frequencies of T cell subsets obtained by flow cytometry was performed using the nonparametric Wilcoxon rank-sum. P < 0.05

## Results

The study included 20 individuals with severe cases of COVID-19 from Lisbon, and 8 individuals with mild to severe cases from Belo Horizonte. The median age of individuals in the infected groups was 69 (IQR 60.0 – 84.3 years) and 42.5 years old (IQR 35.8 – 54.5 years) in Lisbon and Belo Horizonte, respectively. The number of female participants was higher in both control groups when compared to infected patients. The prevalence of comorbidities was higher in the Portuguese population, and hypertension was the most prevalent among infected patients in both cities. Deaths and missing samples were only registered in Lisbon (Table 1).

**Table 1.**
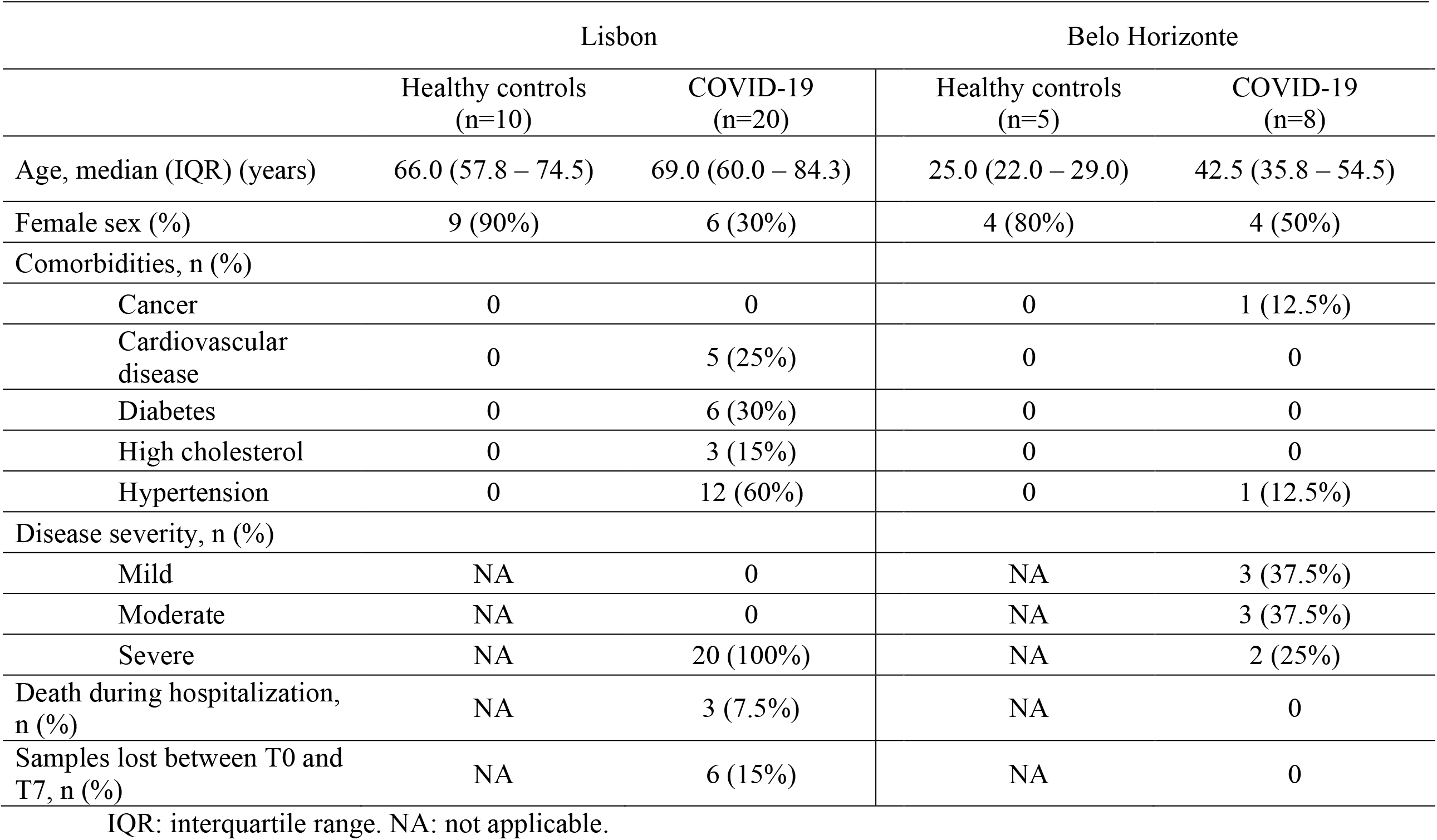
Clinical characteristics of Lisbon and Belo Horizonte populations.

|  | Lisbon |  | Belo Horizonte |  |
| --- | --- | --- | --- | --- |
|  | Healthy controls<br>(n=10) | COVID-19<br>(n=20) | Healthy controls<br>(n=5) | COVID-19<br>(n=8) |
| Age, median (IQR) (years) | 66.0 (57.8 – 74.5) | 69.0 (60.0 – 84.3) | 25.0 (22.0 – 29.0) | 42.5 (35.8 – 54.5) |
| Female sex (%) | 9 (90%) | 6 (30%) | 4 (80%) | 4 (50%) |
| Comorbidities, n (%) |  |  |  |  |
| Cancer | 0 | 0 | 0 | 1 (12.5%) |
| Cardiovascular disease | 0 | 5 (25%) | 0 | 0 |
| Diabetes | 0 | 6 (30%) | 0 | 0 |
| High cholesterol | 0 | 3 (15%) | 0 | 0 |
| Hypertension | 0 | 12 (60%) | 0 | 1 (12.5%) |
| Disease severity, n (%) |  |  |  |  |
| Mild | NA | 0 | NA | 3 (37.5%) |
| Moderate | NA | 0 | NA | 3 (37.5%) |
| Severe | NA | 20 (100%) | NA | 2 (25%) |
| Death during hospitalization, n (%) | NA | 3 (7.5%) | NA | 0 |
| Samples lost between T0 and T7, n (%) | NA | 6 (15%) | NA | 0 |

IQR: interquartile range. NA: not applicable.

### The inflammatory profiles of COVID-19 patients were distinct in the two cohorts

Severe COVID-19 has been reported to be associated with accelerated senescence (Diao et al, 2020; Zheng et al, 2020). In order to understand the immunological profile expressed by the cohorts, we first compared the global production of inflammatory mediators between controls and infected patients in time zero (T0) and time seven (T7). As presented in radar charts (Fig. 1A), there was an overall increase in the frequency of high producers of serum mediators in the infected group from Portugal in T0 when compared to the control group, confirming the ongoing response to the infection. Notably there was an increase in the frequency of high producers of mediators involved in inflammatory response, such as IL-4, IL-6, IFN-γ, TNF, CXCL-5. Seven days after hospitalization, the immune response was still more prominent than in non-infected individuals, despite the reduction in the frequency of high producers of IL-4, CCL-2, IFN-γ and TNF, possibly indicating a tendency towards resolution of the inflammatory response. Corroborating this result, a significant reduction in the production of CCL-2, IFN-γ, IL-6 and IL-10 was also observed (Fig. 1B).

**Figure 1.**
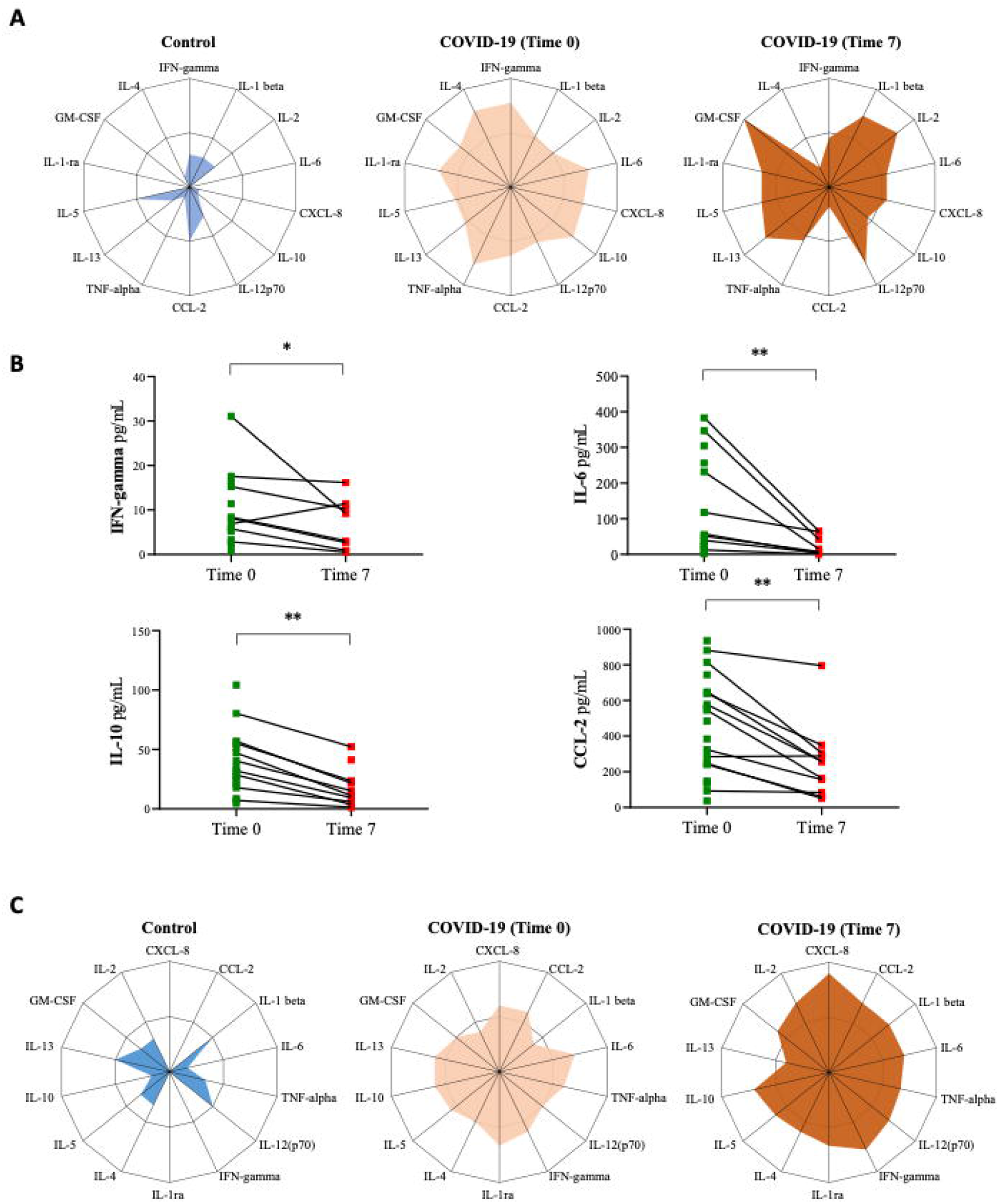
Longitudinal study of inflammatory profiles of COVID-19 patients from Portugal and Brazil during a seven-day period of disease development. (A) Radar chart of serum mediators of control group (baseline) and patients with COVID-19 at hospitalization (T0) and seven days later (T7) in the Portuguese cohort. (B) Comparison of serum levels of CCL-2, IFN-γ, IL-6 and IL-10 at hospital admission (T0) and seven days later (T7). Wilcoxon test was applied to compare samples of the same patient at T0 and T7. P < 0.05. (C) Radar chart representing the balance in frequencies of inflammatory and anti-inflammatory cytokines/chemokines in the innate and adaptive immunity compartments from individuals between the time of recruitment (T0) at the hospital/ambulatory and seven days later (T7) in the Brazilian cohort.

Similar to the analysis conducted for Lisbon patients, to assess whether a population composed of younger individuals from Brazil, who developed moderate to severe clinical conditions, presented a similar pattern of longitudinal immunological response, we compared the overall inflammatory profile of control and infected patients (Fig. 1C). As expected, the control group presented a very discreet production of mediators, while infected individuals showed a burst in inflammation throughout the study. When comparing T0 and T7, there was a rise in the frequency of high producers of many cytokines, including CXCL-8, IL-10, IL-2, GM-CSF, IL-4, IFN-γ, and IL-12p70, indicating intensification in the inflammatory response. Meanwhile, IL-13 presented a reduction in its frequency (Fig 1C). No significant change in the levels of these mediators was observed within seven days.

Longitudinal correlation analysis of immunological mediators allowed us to gain insights into cytokines, chemokines, and growth factors production during hospitalization (Fig. 2A). In the Lisbon cohort, although we observed a reduction in the number and intensity of correlations, there was a remarkable switch from positive to negative interactions between mediators, notably in CXCL-2, CCL-2, IL-6, IL-12p70, INF-γ, and IL-4. Some mediators, such as IL-2, GM-CSF and IL-13 gained interactions in T7 (Fig. 2A).

**Figure 2.**
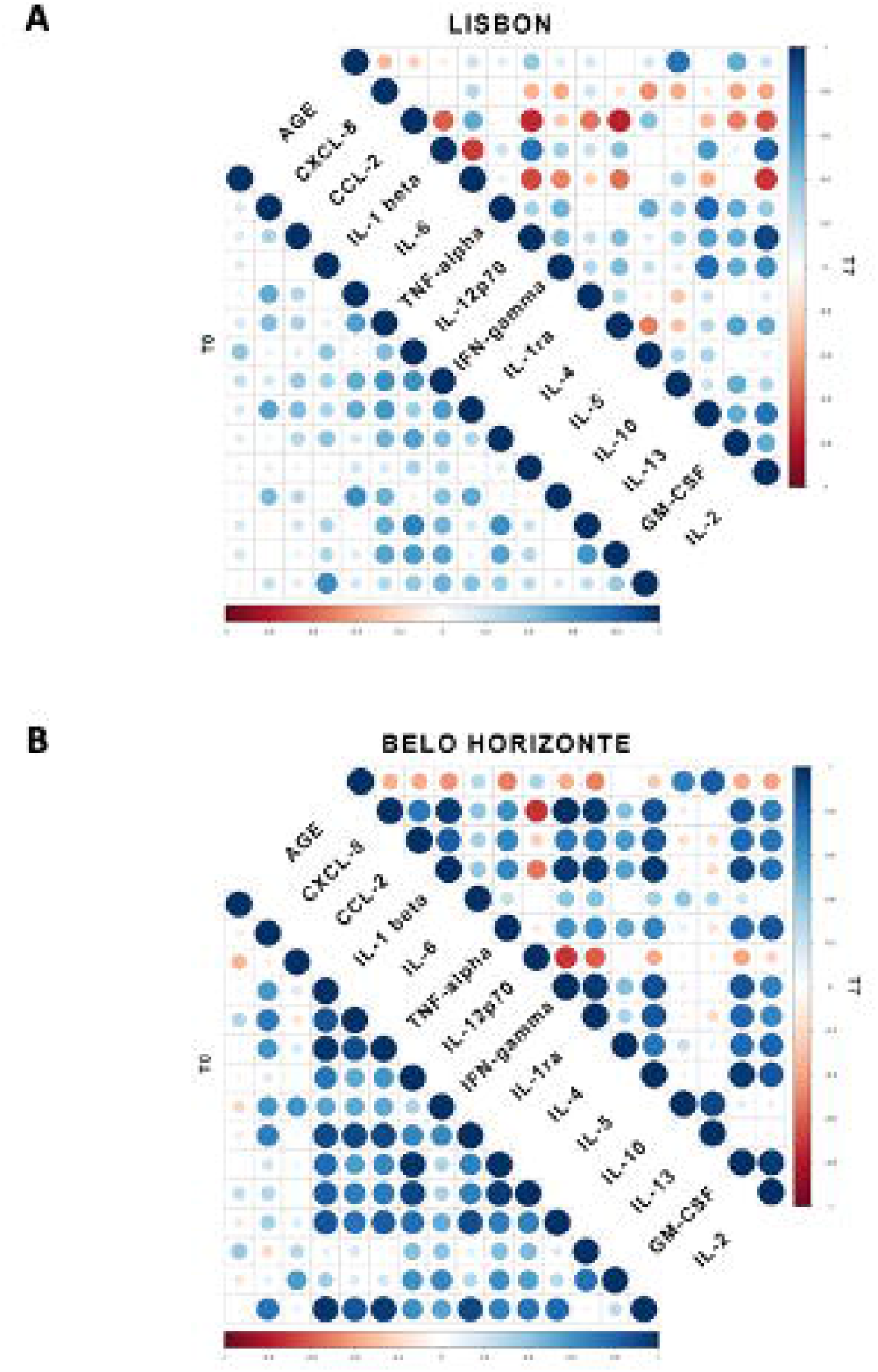
Spearman’s correlation correlogram of the inflammatory profile of COVID-19 patients at day 0 and 7 of disease development. (A) Correlation matrices across all cytokines/chemokines from patients’ sera, comparing COVID-19 at time zero (T0) of infection and 7 days later (T7) in Lisbon, Portugal, and (B) Belo Horizonte, Brazil. Positive cytokine correlations are lost, and negative correlations appear going from T0 to T7. The strength of the correlation between two variables is represented by the color of the circle at the intersection of those variables. Colors range from dark blue (strong positive correlation, i.e. r2 = 1.0) to dark red (strong negative correlation; i.e. r2 = -1.0). Results were not displayed if not significant (P < 0.05) according to Spearman Correlation Test.

The correlation matrix graphic representation from Brazil showed a decline in some interactions within seven days. There is an evident loss of positive correlations for IL-6, IL-10, TNF, and IL-12p70, with the latter having also gained negative correlations, notably with CXCL-8, IL-β, IL-1ra, and INF-γ (Fig. 2C). On the other hand, CCL-2, IFN-γ, and GM-CSF gained intensity and significance in numerous interactions (Fig. 2C). Interestingly, we see an increase in the total number of significant correlations between age and cytokines, mostly negative ones, which could indicate a reduction in the interactions with aging.

### Exhausted and immunosenescent T cells accumulate in COVID-19 patients of both cohorts regardless of disease severity

Since inflammatory response during COVID-19 infection is already associated with T cell exhaustion (Arcanjo et al, 2021; Lee et al, 2021), we used flow cytometry immunophenotyping to evaluate alterations in immune responses throughout the course of infection. We investigated the evolution of the relative abundance of CD4^+^ and CD8^+^ T cells using manual gating as described in Supplementary Figure 1. T cell markers related to differentiation (CD28, CCR7, CD45RO) and senescence/exhaustion (CD57, KLRG1, PD-1) allowed us to identify these phenotypes in distinct cell compartments including naïve (CD28^+^CCR7^+^CD45RO^-^) and effector memory (CD28^-^CCR7^-^ CD45RO^+^) T cells.

Results from Lisbon patients showed an increase in the frequency of differentiated T CD4^+^ cells (CD28^-^) expressing both markers of senescence/exhaustion (CD57^+^KLRG1^+^) as well as effect memory cells (EM) (CD45RO+ CCR7-CD28-) (Fig. 3 C and E). In the CD28-CD8^+^ T cell compartment, frequency CD57^+^KLRG1^+^ cells increased while there was no difference in EM subset (Fig. 3 D and F).

**Figure 3.**
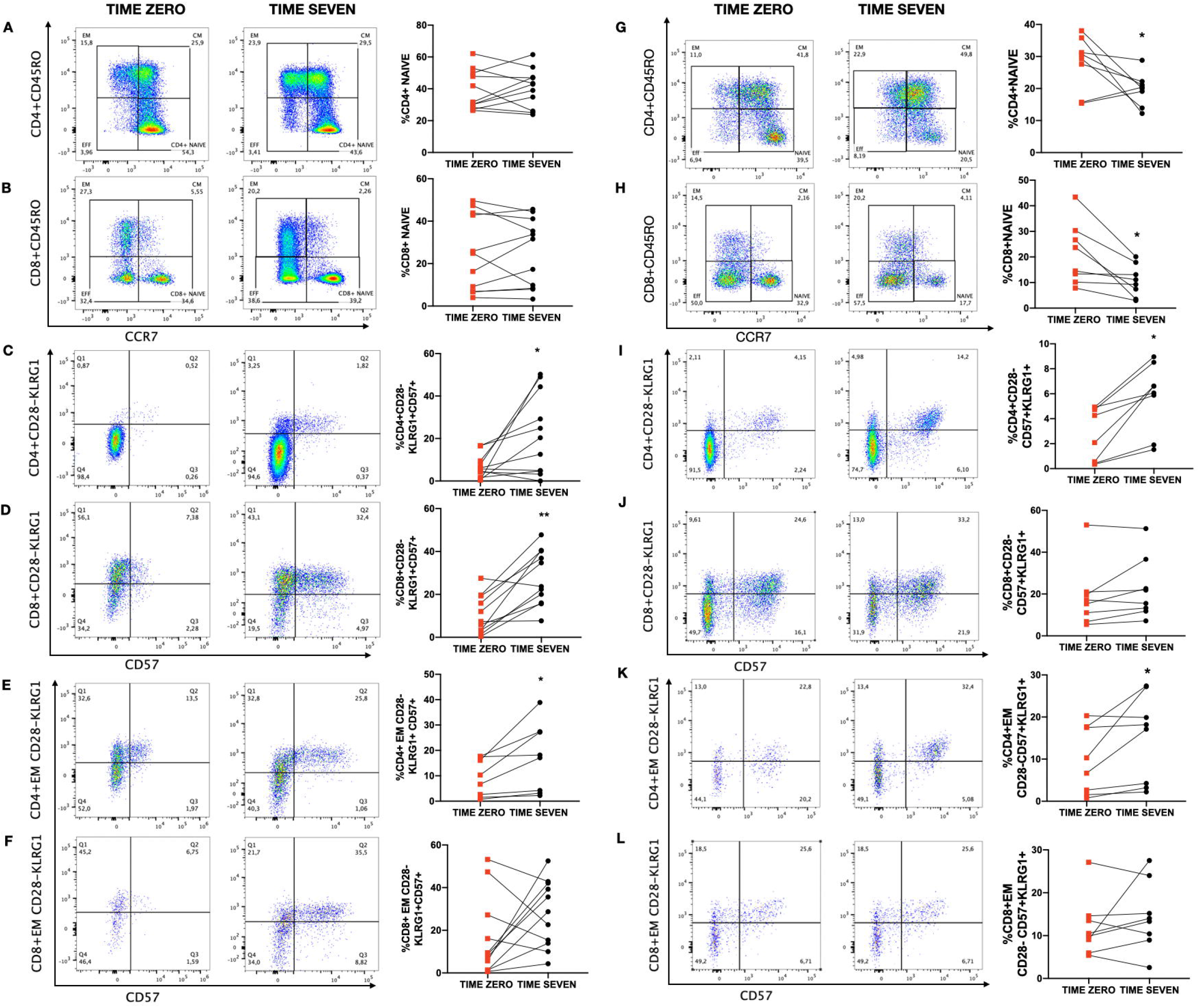
Frequencies of exhausted/senescent T cells in patients with mild to severe COVID-19 from Portugal and Brazil during a seven-day period. Representative flow cytometry plots of gated cell subsets and frequency analysis at time 0 and 7. (A and B) Frequencies of naïve CD4 and CD8 T cell in individuals from Lisbon. (C, D, E and F) Differentiated CD4+ and CD8+ T cells presenting senescence/exhaustion markers CD57 and KRLG1 obtained by manual gating in Portuguese patients. (G-L) Percentage of CD4+ and CD8+CD28-CD45RO+CCR7-(effector memory) expressing senescence/exhaustion markers in Brazilian patients. Statistical analysis was performed using the nonparametric Wilcoxon rank-sum. P < 0.05.

Contrary to what was observed in severe cases from Lisbon, frequencies of naïve CD4^+^ and CD8^+^ T cells from patients in Brazil reduced during hospitalization (Fig 3 G and H). This change was accompanied by an increase in T CD4^+^CD28^-^ and EM cells double-positive for CD57^+^KLRG1^+^ during seven days (Fig. 3 I and K). It was also observed a reduction in the frequency of FOXP3^+^ CD4^+^ T cells in the individuals from the Brazilian cohort whereas no difference was observed in the frequency of PD-1-expressing (exhausted) FOXP3^+^ CD4^+^ T (Fig S2).

Altogether our results show an emergence of an increased senescent/exhausted T cell phenotype in patients with COVID-19 during 7 days of infection regardless of disease severity.

## Discussion

Inflammation induced by viral infections plays an important role in the differentiation and development of T cells, as they induce extensive cell proliferation, however it can ultimately lead to cell exhaustion and senescence (Wherry, 2011; Moro-Garcia et al, 2018; Heath & Grant, 2020). Although this phenomenon is mainly related to chronic infections such as CMV, HIV, HBV and HPV (Fulop, Larbic, Pawelec, 2013; Wherry, 2011), it has also been identified in patients with acute SARS-Cov-2 infection (De Biase et al, 2020; Arcanjo et al, 2021). Considering these findings and the highly heterogeneous immunological responses observed in COVID-19 (Lucas et al, 2020), we studied the evolution of the inflammatory response and exhaustion/senescence T cell profile in a seven-day interval in two different cohorts of infected individuals (one from Belo Horizonte, Brazil and another one from Lisbon, Portugal).

Our longitudinal analysis revealed different inflammatory profiles between cohorts, although both presented a hyperactivation of the immune system in response to the infection. At admission, the severe patients from Portugal exhibited high levels of biomarkers associated with a core response signature in moderate to severe conditions in COVID-19, such as IFN-γ, TNF, IL-6, and IL-10 (Liu et al, 2020; Lucas et al, 2020). Meanwhile, over the seven-day period, there was a trend towards the reduction of the inflammatory profile, as observed by the decrease in all cytokine levels (Fig. 1A). This data is supported by the significant reduction of IL-2, IFN-γ, and IL-6 (Fig. 1B), which are biomarkers related to hyperactivation of T cells and recruitment of macrophages to the site of infection (Arcanjo et al, 2021; Coperchini et al, 2021). Decrease in the anti-inflammatory cytokine IL-10 accompanies the general decline in inflammatory immune response. The emergence of negative correlations in the correlation matrix also suggests the downregulation of the immune response during the course of the disease (Fig. 2B), indicating a tendency to resolution of the inflammatory response.

Patients from Brazil with mild to severe disease had a trend towards a global rise in serum cytokine production (Fig. 1C). For instance, the frequency of high producers of CXCL-8, a chemokine involved in the recruitment of neutrophils to the site of infection and commonly elevated in patients with COVID-19 (Coperchini et al, 2020; Sallenave & Guillot, 2020; Hazeldine & Lord, 2021), suggests an enhancement of the inflammatory response in the course of seven days (Fig 1C). Also, the correlation matrix evidenced higher correlations between CCL-2, IFN-γ, and GM-CSF levels in T7. These mediators are implicated in T lymphocyte response to SARS-CoV-2, such as cell trafficking, cytokine production and cell replication (Coperchini et al, 2020; Arcanjo et al, 2021). Considering that disease is associated with the loss of complexity and dynamics of physiological processes (Lipsitz, 2004), the reduction in some correlations after a week indicates a putative imbalance in the immune network complexity developed during this short period of infection (Fig 2A).

As a complement to the inflammatory profile found in COVID-19 patients from Brazil, we also observed a reduction in the frequency of Foxp3^+^CD4^+^ T cells which have been widely described as regulatory cells mediating anti-inflammatory effects. Of note, frequency of Foxp3^+^CD4^+^ T cells expressing the exhaustion marker PD-1 did not change suggesting that only functionally active regulatory T cells were affected by the infection. These data are in accordance with previous reports showing a reduction in the frequency of these T cell subset in patients with COVID-19 namely in patients with a severe form of the disease (Lucas et al, 2020).

It is possible that the two cohorts, in Brazil and Lisbon, were not exactly synchronized. In fact, we observed a more marked reduction of pro-inflammatory cytokines between day 0 and 7 in the Lisbon cohort. However, although this heterogeneity may sound as a negative feature of our sample, the similar results yielded by the two distinct cohorts regarding the immunosenescence profile of the T cell compartment helped to reveal a common phenomenon associated with SARS-CoV2 infection.

Studies on individuals with severe COVID-19 revealed an increase in T cells expressing markers of exhaustion/senescence such as PD-1, TIGIT, and CD57, especially in the CD8 compartment, when compared to healthy individuals (Zheng et al., 2020a; Zheng et al, 2020b; De Biasi, 2020; Arcanjo et al, 2021). In the present work, our aim was to examine the emergence of these alterations in a longitudinal study and to determine whether these immunosenescence alterations were restricted to severe cases of the disease in two distinct cohorts. Our data show that patients with mild and severe cases of COVID-19 in Brazil had an increase in CD4^+^CD28^-^ T cells expressing both CD57 and KLRG1, whereas patients from Lisbon with severe disease showed this increase in both CD4^+^CD28^-^ and CD8^+^CD28^-^ T cell compartments (Fig 3 C, D, I). Expression of CD57 and KLRG1, and the loss of the CD28 molecule are considered indicators of terminal differentiation and are associated with the senescence profile of T lymphocytes (Akbar & Henson, 2011; Pangrazzi & Weinberger, 2020). Although the KLRG1 molecule also emerges on the cell surface after differentiation, the signalling generated by this molecule regulates a pathway related to exhaustion, making it a marker of cellular senescence and exhaustion (Akbar & Hsenson, 2011; Wherry & Kurachi, 2015; Pinti et al, 2016; Cunha et al, 2020; Pangrazzi & Weinberger, 2020). Therefore, absence of CD28 and the high expression of CD57 as well as KLRG1 provide what could be considered the best description of a senescent state (Rodriguez et al, 2021).

Interestingly, our results identified the emergence of a senescence profile of T lymphocytes in individuals with COVID-19, both in mild/moderate and severe conditions, being more pronounced the more intense the inflammatory response to the disease was. Although there is no data in the literature relating inflammation and immunosenescence in COVID-19, the hyperactivation of the inflammatory response to SARS-CoV-2 infection has been associated with more severe disease (Lucas et al, 2020), which could possibly justify the increase in the frequency of terminally differentiated cells in this group of patients.

It is also important to highlight that the cohort of critically ill patients from Portugal is mainly composed of elderly individuals. Thus, this may explain the difference between health conditions between Brazilian and Portuguese cohort samples. As predicted by our group and by others, immunosenescence and inflammaging may be aggravated by, but also may aggravate SARS-CoV-2 infection (Batista et al, 2020; Witkowski, Fulop, Bryl, 2022). The primary setting and one of the hallmarks of immunosenescence is thymic involution, which is followed by the reduction in the naïve cell output (Thomas, Wang, Su, 2020; Elyahu & Monsonego, 2021), a phenomenon that should have been affecting these patients even before the onset of the infection. This could be the reason why no difference was observed in the naïve T cell subset in this severely diseased elder group.

The differences seen in inflammatory biomarkers between patients of Brazil and Portugal may be related to the time of the onset of symptoms, as the recruitment was carried out based on the moment patients sought medical assistance at the hospital, rather than on the initiation of symptoms. Notably, accumulation of exhausted/senescent cells seems to occur at distinct time points of disease development, indicating a process that could start early in infection and develop further than the 7-day period examined, particularly in Brazil where the patients were recruited sooner regarding the course of the disease.

There were limitations in this study, mainly the small sample size in both cohorts. This may have hindered the identification of significant differences in inflammatory mediators and other T-cell subsets that would be possible using a larger number of individuals. Moreover, this feature precluded formal statistical analysis regarding the associations between laboratory findings and disease severity. There were also methodological differences in the study of the two cohorts that could have impacted on the results. We did not investigate the SARS-CoV-2 variant infecting the individuals and this could also partially explain differences in their immunological responses, considering the distinct scenarios of the pandemic in Brazil and Portugal during which patients were recruited.

Our longitudinal study provided evidence of accelerated immunosenescence in the T cell compartment during COVID-19 showing that senescent and exhausted T cells significantly increased within a seven-day period. Interestingly, even though our samples were composed by two different populations, who presented distinct inflammatory profiles and were possibly affected by different variants of the virus, a similar profile of immunossenescence and exhaustion was identified during this short period of infection by SARS-CoV-2. Furthermore, this change in immunological profile was shown to be independent of disease severity even though the magnitude was greater in patients with severe disease. This suggests that accelerated immunosenescence of the T cell compartment may be described as a general feature of COVID-19.

## Data Availability

All data produced in the present study are available upon reasonable request to the authors

## Acknowledgments

This study was financially supported by grants from Fundação para a Ciência e a Tecnologia (FCT, Portugal, Research4covid_369), Merck Sharp & Dohme (MISP 42521) and Conselho Nacional de Desenvolvimento Científico e Tecnológico (CNPq, Brazil, APQ 407363/2021-1). RB Pedroso is funded by FCT (SFRH/BD/144372/2019). Samples in Lisbon were collected, stored and provided by Biobanco-iMM (Lisbon Academic Medical Center, Lisbon, Portugal) in coordination with a taskforce created at iMM for the collection of COVID-19 Research Samples. Flow cytometry in Lisbon was performed at the Flow Cytometry Facility of iMM.

## Conflict of interest

The authors have no conflict of interest.

## Figure Legends

**Figure S1 - Gating strategy used to identify the profile of exhausted and senescent CD8+ and CD4+ T cells.** (A) T cells were characterized based on their CD3 and CD4 expression, and then categorized by CD45RO, CCR7, CD28, CD57 and KLRG1 expression. Regulatory T cells expressing FOXP3 were assessed in Brazilian population for their PD-1 expression. (B) CD8 Gate strategy for analysis of CD8+ T cells was similar to the one used for CD4+ T cells.

**Figure S2 - SARS-CoV-2 infection decreases the frequency of FOXP3+CD4+ T cells in Brazilian patients with mild to severe COVID-19 in a seven-day period.** Percentage of T CD4+CD25+FOXP3+ cells obtained by manual gating. Frequency of cells expressing exhaustion marker PD-1. Statistical analysis was performed using the nonparametric Wilcoxon rank-sum. P < 0.05

## Notes

### Competing Interest Statement

The authors have declared no competing interest.

### Funding Statement

This study was financially supported by grants from Fundacao para a Ciencia e a Tecnologia (FCT, Portugal, Research4covid_369), Merck Sharp & Dohme (MISP 42521) and Conselho Nacional de Desenvolvimento Cientifico e Tecnologico (CNPq, Brazil, APQ 407363/2021-1) RB Pedroso is funded by FCT (SFRH/BD/144372/2019)

### Author Declarations

Samples in Lisbon were collected, stored and provided by Biobanco iMM (Lisbon Academic Medical Center, Lisbon, Portugal) in coordination with a taskforce created at iMM for the collection of COVID 19 Research Samples Flow cytometry in Lisbon was performed at the Flow Cytometry Facility of iMM The research protocol was approved by the National Research Ethics Commission (CONEP # 5.190.260) of Brazil and by the Lisbon Academic Medical Center Ethics Committee (ref. no. 306/20). The study was conducted in accordance with the Helsinki Declaration for research involving humans. All participants, including healthy controls, agreed to participate voluntarily in this study without financial support and signed an informed consent.

