## Supplementary Table 1 for "COVID-19 INDUCES SENESCENCE AND EXHAUSTION OF T CELLS IN PATIENTS WITH MILD/MODERATE AND SEVERE DISEASE DURING A SEVEN-DAY INTERVAL"

**Supplementary Table 1 –** Biomarkers measured by Multiplex (BioRad Bio-Plex Pro Human Cytokine Standard) and their standard settings

| ANALYTE | DESCRIPTION | Standard PMT Setting (pg/mL) |
|---|---|---|
| **IL-1β** | Interleukin – 1 beta | 1,013 |
| **IL-1Ra** | Interleukin – 1 alpha | 76,896 |
| **IL-2** | Interleukin – 2 | 17,225 |
| **IL-4** | Interleukin – 3 | 1,588 |
| **IL-5** | Interleukin – 5 | 57,568 |
| **IL-6** | Interleukin – 6 | 7,980 |
| **CXCL8** | Chemokine Ligand-8 | 17,258 |
| **IL-10** | Interleukin – 10 | 26,569 |
| **IL-12p70** | Interleukin – 12p70 | 19,101 |
| **IL-13** | Interleukin – 13 | 4,658 |
| **GM-CSF** | Granulocyte-Macrophage Colony-Stimulating Factor | 6,627 |
| **IFN-γ** | Interferon-gamma | 4,495 |
| **CCL2** | Chemokine Ligand-2 | 7,634 |
| **TNF-α** | Tumoral Necrosis Factor-alpha | 53,044 |
