## Supplementary figures and images for "COVID-19 INDUCES SENESCENCE AND EXHAUSTION OF T CELLS IN PATIENTS WITH MILD/MODERATE AND SEVERE DISEASE DURING A SEVEN-DAY INTERVAL"

### Supplementary Figure 1

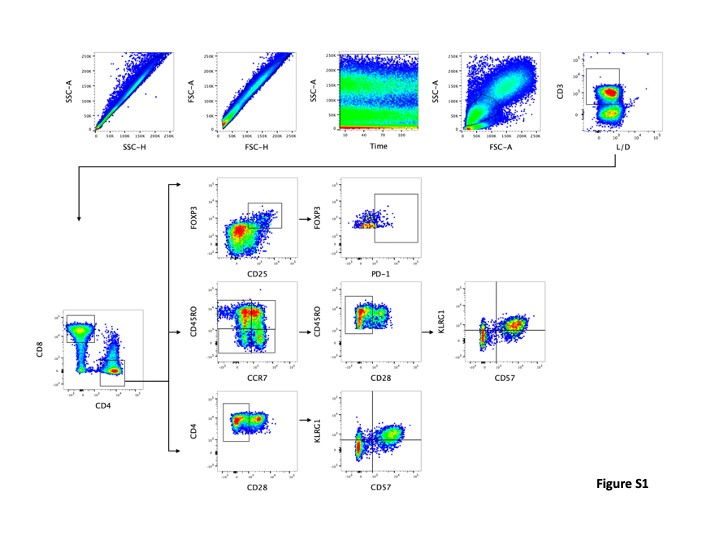

### Supplementary Figure 2

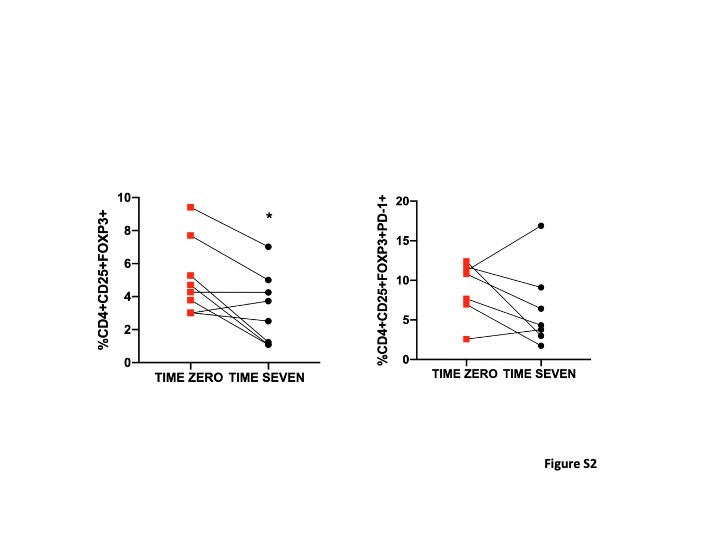
